# Tracking SARS-CoV-2 seropositivity in rural communities using blood-fed mosquitoes

**DOI:** 10.1101/2023.06.13.23291267

**Authors:** Benjamin J. Krajacich, Djibril Samaké, Adama Dao, Moussa Diallo, Zana Lamissa Sanogo, Alpha Seydou Yaro, Amatigué Ziguimé, Josué Poudiougo, Kadiatou Cissé, Mamadou Traoré, Alassane dit Assitoun, Roy Faiman, Irfan Zaidi, Woodford John, Patrick Duffy, Tovi Lehmann

**Affiliations:** Laboratory of Malaria and Vector Research, NIAID, NIH, Rockville, Maryland, USA; Malaria Research and Training Center (MRTC)/ Faculty of Medicine, Pharmacy and Odonto-stomatology, University of Sciences, Techniques and Technologies, Bamako, Mali; Laboratory of Malaria Immunology and Vaccinology, National Institute of Allergy and Infectious Diseases, National Institutes of Health, Bethesda, MD, USA

**Keywords:** Africa, COVID-19, population-based epidemiology, mosquito blood meal, sero-surveillance

## Abstract

The spread of SARS-CoV-2 cannot be well monitored and understood in areas without capacity for effective disease surveillance. Countries with a young population will have disproportionately large numbers of asymptomatic or pauci-symptomatic infections, further hindering detection of infection in the population. Sero-surveillance on a country-wide scale by trained medical professionals may be limited in scope in resource limited setting such as Mali. Novel ways of broadly sampling the human population in a non-invasive method would allow for large-scale surveillance at a reduced cost. Here we evaluate the collection of naturally bloodfed mosquitoes to test for human anti-SARS-CoV-2 antibodies in the laboratory and at five field locations in Mali. Immunoglobulin-G antibodies were found to be readily detectable within the mosquito bloodmeals by a bead-based immunoassay at least through 10 hours post-feeding with high sensitivity (0.900 ± 0.059) and specificity (0.924 ± 0.080), respectively, indicating that most blood-fed mosquitoes collected indoors during early morning hours (and thus, have likely fed the previous night) are viable samples for analysis. We find that reactivity to four SARS-CoV-2 antigens rose during the pandemic from pre-pandemic levels. Consistent with other sero-surveillance studies in Mali, crude seropositivity of blood sampled via mosquitoes was 6.3% in October/November 2020 over all sites, and increased to 25.1% overall, with the town closest to Bamako reaching 46.7% in February of 2021. Mosquito bloodmeals a viable target for conventional immunoassays, and therefore country-wide sero-surveillance of human diseases (both vector-borne and non-vector-borne) is attainable in areas where human-biting mosquitoes are common, and is an informative, cost-effective, non-invasive sampling option.

## Introduction

The speed, scope, and impact of the SARS-CoV-2 virus on all corners of the globe is unprecedented in the last 100 years, with over 662 million cases and 6.7 million deaths through 2022.^1^ As of February 2022, Mali had 30,303 RT-PCR-confirmed cases of COVID-19 across four waves of infection for a population of 20.8 million (Figure 1)^2^, with most cases reported from the capital, Bamako. Due to the limited testing capacity across the country, this is almost certainly a gross underestimation of the true number of infections. The use of sero-surveillance in which blood samples are broadly screened for anti-SARS-CoV-2 antibodies, is a promising tool to discover the rate of spread of a disease through a population even from individuals without known current or past infection.

**Figure 1:**
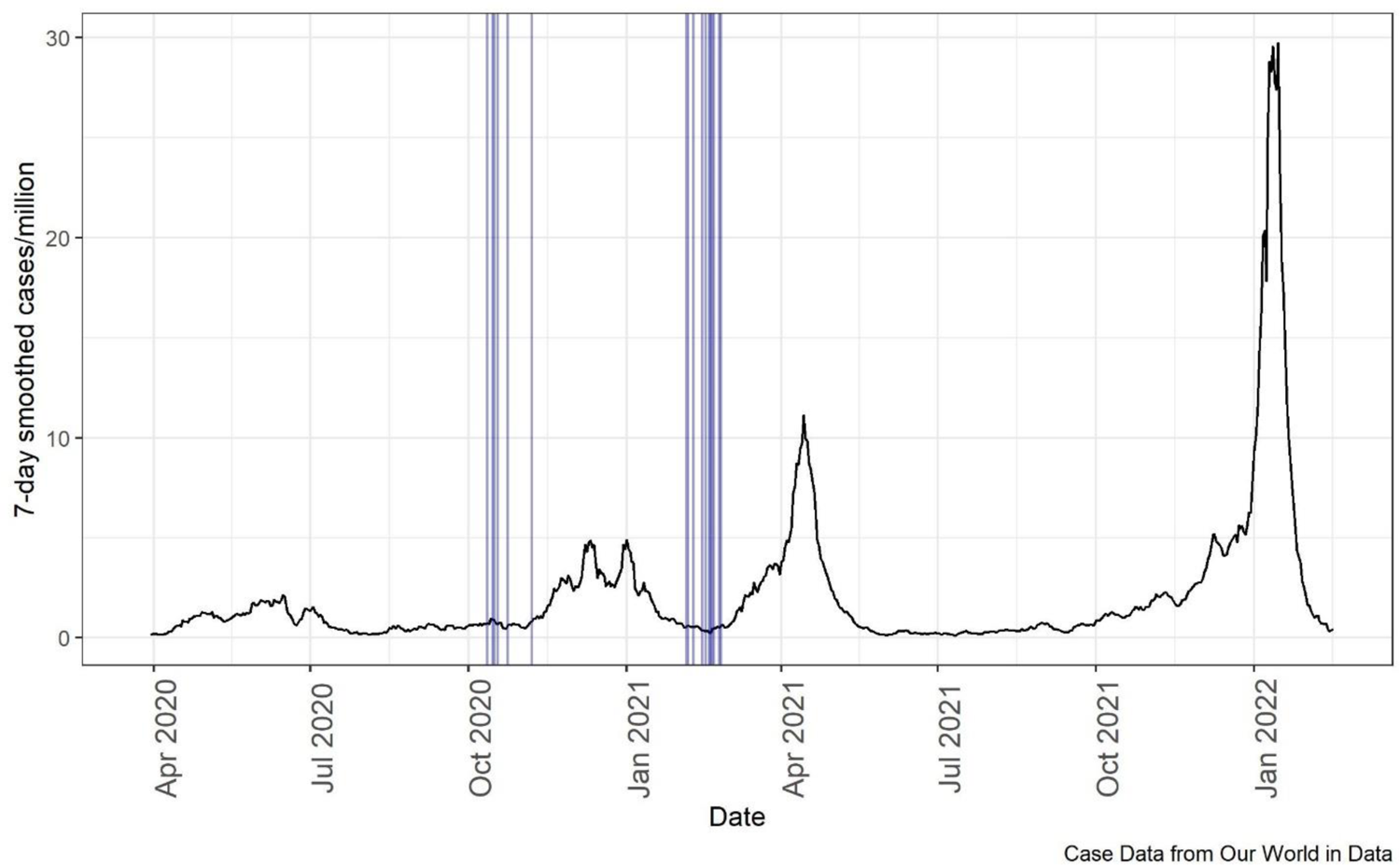
SARS-CoV-2 7-day smoothed case numbers per million people in Mali. Mosquito sampling dates across all villages marked by vertical blue lines. Data from Our World In Data.^1, 2^ As of 2022-02-17, 30303 cases total in population of 20.8 million.

Enzyme-linked immunosorbence assays (ELISAs) for detection of immunoglobulin-G antibodies specific for SARS-CoV-2 have been rapidly developed in response to the pandemic.^3–5^ However, samples of African origin tested with these assays were associated with a higher background reactivity than has been seen with North American and European serum panels, though this varied depending on the assay used and antigen targeted.^6–8^ The origins of the background reactivity have not been fully explained, though donors in some of the African panels had higher reactivity to ‘common-cold’ coronaviruses such as OC43^6^ and previous infections with Dengue virus or malaria-causing parasites may also exacerbate this problem^9^. Within Mali, background reactivity in pre-pandemic sera was common compared to US controls (43.4% spike, 22.8% for receptor-binding domain (RBD), and 33.9% for nucleocapsid protein), with no obvious neutralization ability present in these samples.^8^ Specificity could be improved with reasonable sensitivity by using multiple antigen targets with conservative cutoffs, though the degree to which this background signal may impact other immunoassays is unclear.

Previous work with this dual-antigen ELISA method in Mali has indicated a sharp increase in the adjusted seroprevalence from 10.9% in July-October 2020 to 54.7% in December 2020-January 2021 in three villages/townships.^10^ This absolute increase appeared to be more limited in the relatively rural village of Doneguebougou (from 5.0 to 37.0%), in comparison to the Sotuba township nearby to Bamako (from 19.0 to 70.4%), but was still dramatic. Although sampling in this study included the township of Bancoumana, 50 km from Bamako, it was not performed in rural areas further from the capital, so the representation of the country and the relationship between distance from Bamako and rate of seropositivity remained unclear. Expansion of sero-surveillance into more remote and rural areas of Mali may be difficult due to requirements of trained health professionals for blood-collection and analysis.

Reflecting on our experience in medical entomology, we considered the blood sampling potential present in human-seeking mosquitoes. In many areas of Mali, the mosquito density indoors is high, especially of a few species such as certain members the *Anopheles gambiae* and *Culex pipiens* complexes, which have a strong preference for blood-feeding from humans, and a tendency to rest indoors after imbibing a bloodmeal.^11^ With these numbers and behaviors, they present a suitable potential source for an assay-sized, roughly 1-5 µl^12^, volume of blood. Previous work in this realm has found mosquito bloodmeals to have sufficient volume to detect blood-borne human pathogens,^13^ and various antibodies of human disease including *Trypanosoma, Plasmodium,* Dengue virus, and Japanese Encephalitis antibodies.^14–16^ However, leveraging these insects has never, to our knowledge, been performed as a broad epidemiological sero-surveillance tool.

In this study we evaluate the potential for anthropophilic biting mosquitoes as non-invasive blood sampling tools to measure population seroprevalence patterns, using naturally acquired SARS-CoV-2 antibodies as a proof of concept. Owing to a relatively small amount of blood drawn via a mosquito bite, we favored a multiplex, bead-based immunological assay to characterize four antigens simultaneously. We characterize the durability of antibodies in the digestive environment of the mosquito midgut and evaluate the methodology in natural settings using mosquitoes caught in five different communities in Mali, West Africa.

## Methods

### Mosquito rearing

*Anopheles coluzzii* mosquitoes were reared as previously described.^17^ Briefly, colonized, disease-free mosquitoes were reared in plastic trays (30 x 25 x 7 cm) with 1.5L of dechlorinated water. Larvae were fed with yeast supplement in the first 24 hours post after larval emergence and fish food until emergence as adults. These adult mosquitoes were held until 3-5 days old, at which point they were starved overnight and allowed to feed upon human volunteers and stored as described below.

### Pre-pandemic mosquito collection and multiplex bead-based immunoassay reactivity

To develop a baseline cutoff from historic samples, bloodfed mosquitoes collected as part of mosquito surveillance from the Sahelian villages Thierola and M’Piabougou, Mali in 2017 and 2018, that had been stored on silica gel desiccant were evaluated for immunoreactivity to SARS-CoV-2 antigens. Abdomens of individual mosquitoes were separated from the thorax under magnification using fine-tipped forceps. These abdomens were ground individually in 120ul of sample buffer of the bead-based kit (Bio-Plex Pro Human IgG SARS-CoV-2, Bio-Rad, Irvine, CA) using 5-6 2.0 mm zirconia beads in a Mini-BeadBeater-96 (BioSpec Products, Inc, Bartlesville, OK, USA) at max speed for 25 seconds. This slurry was spun at 13,000 g for 10 minutes to clear solids, and 50ul of the supernatant was used in the assay according to manufacturer instructions. This assay uses anti-human IgG as detection antibody and magnetic capture beads coupled with SARS-CoV-2 nucleocapsid, receptor-binding domain, spike subunit 1 (hereafter spike1), and spike subunit 2 (hereafter spike2) viral proteins. Volume of sera per bloodmeal is difficult to quantify with the mosquito due to variability in feeding amounts and the concentration of blood via diuresis. For each antigen, we developed cutoffs from pre-pandemic samples based on the time period’s mean value of sample median fluorescence intensity (MFI) plus 5 standard deviations, and samples were considered positive if two or more antigens exceeded these cutoffs.

### Human blood feeding on SARS-CoV-2 recovered individuals and computation of sensitivity and specificity

#### Disclaimer

All aspects of the work involving human volunteers were approved by the Ethics Committee in the University of Bamako as part of IRB protocol: N°2020/78/CE/FMOS/FAPH.

Two volunteers who had recovered from a PCR-diagnosed SARS-CoV-2 infection, and one volunteer with no known SARS-CoV-2 infection were fed upon by groups of 100 laboratory-reared, disease-free *An. coluzzii.* Mosquitoes were kept under normal insectary conditions for set time points post feed (0, 2, 4, 8, and 30 hours) to analyze the effect of digestion on recoverability of the antibodies. At each time point, mosquitoes were killed and stored on top of a small piece of cotton ball in a 1.7 ml Eppendorf tubes with silica gel desiccant (#13767, Millipore Sigma, Burlington, MA). Mosquitoes were kept on desiccant for one week at room temperature before storage at −80 °C until analysis. A larger cohort of 12 residents of the rural village of Thierola of unknown SARS-CoV-2 status were subsequently analyzed. Of this cohort, two (C and K) had worked in a mine during several months prior to the experiment and two lived in cities (‘H’ in Bamako and ‘M’ in Kita). An additional volunteer, a resident of Bamako, was vaccinated against SARS-CoV-2 three months prior to the experiment. Each volunteer was fed upon by groups of 50 laboratory-reared, disease-free *An. coluzzii* mosquitoes. Mosquitoes were held for 0, 5, 10, and 30 hours post-feed and stored as described above before analysis. Using the ‘rsample’ R package^18^, we created 100 sample splits for each timepoint using bootstrapping. From these splits, we estimated test sensitivity and specificity from the putative positivity of the volunteers using with the ‘yardstick’ R package.^19^ Overall test sensitivity and specificity were set based on 5 and 10 hr timepoints as these are the most-likely range of time periods post-feeding to capture mosquitoes (i.e. morning after likely feeding window).

### Wild-caught mosquito based sero-surveillance in Mali and evaluation of changes in crude prevalence

Indoor resting mosquitoes were collected by aspiration from 40 sentinel houses in each of five communities (latitude and longitude are in parentheses): Bancoumana (12.20862, −8.2646), Berian (11.4197, −7.9351), Nionina (12.9873, −5.997231), Sitokoto (13.637307, −10.818615), and Sotuba (12.66181, −7.91915), Mali. The sentinel houses were chosen to have at least one occupied bedroom, to have given permission for sampling via the homeowners, and to be spread across the community with a minimum of 50 m between them (20 m in the smaller villages). Sampling of blood-fed mosquitoes was performed during October/November 2020 and February 2021 with handheld aspirators in the morning (07:00-10:00). Mosquitoes were desiccated on silica gel and stored at −20 °C until shipment where they were stored at −80 °C until analysis. Samples of ∼60 blood-fed mosquitoes per village per timepoint were analyzed via the below described immunoassay. Where possible, mosquitoes were sampled from the same houses across time periods to assess the progression of seropositivity in a semi-matched population. These are semi-matched due to an uncertainty whether the individual the mosquito fed on is a member of that house, and if so which individual it was.

To test the difference between quantiles of pre-pandemic and pandemic distributions, we used quantile regression implemented by Proc Quantreg^20^, which extends the general linear model for estimating conditional change in the response variable across its distribution as expressed by quantiles, rather than its mean (though the median is similar to the mean in symmetric distributions). Quantile regression does not assume parametric distribution (e.g., normal) of the random error part of the model, thus it is considered semi-parametric. The benefit of this analysis is that it addresses changes in reactivity to antigens that could be detected in the higher quantiles even when the mean or the median are less affected, without imposing cutoffs. In fact, it can be used to estimate if there is a monotonic increase over time (and over quantiles) and estimate the sero-prevalence change (from the pre-pandemic baseline) per antigen. The parameter estimates in linear quantile regression models are interpreted as in typical GLM, as rates of change adjusted for the effects of the other variables in the model for a specified quantile^21^.

### Seropositivity adjustments

The definition of seropositive blood meal (hereafter also seropositive mosquito) is explained in the Results (below). Estimated seropositivity at the human population was adjusted in three ways, to account the assay sensitivity and specificity (calculated from 5hr and 10hr time-points of laboratory feed), for human-blood meal proportion, and to account for the possibility that multiple mosquitoes fed on the same person. Specifically, standard error around crude seropositivity measures per village per timepoint were generated using the ‘yardstick’, ‘infer’, and ‘rsample’ packages^18, 19, 22^ with 1,000 bootstrap replicates. The test sensitivity and specificity adjustment was done as in Sempos and Tian (2021)^23^ using the sensitivity and specificity estimates from the 5hr and 10hr time-points of laboratory feed, with the following formula:

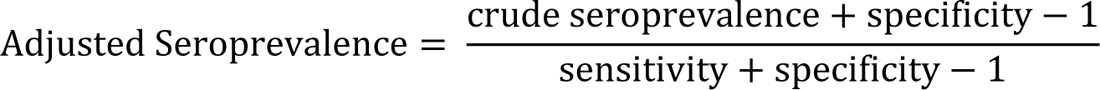

Second, we adjusted seroprevalence estimates accounting for the non-human bloodmeal proportion found in mosquito samples collected during the matched village and time periods (methodology described in section below), adjusting the numbers of total individuals tested (denominator) by the corresponding fraction of non-human bloodmeals found. Uncertainty around these estimates were taken by generating 1000 draws stratified by sample group size, and 1000 bootstrap replicate from these groups as above. Finally, seroprevalence when sampling a single mosquito per house was estimated across 1,000 random draws of one mosquito per house per time period. Standard errors surrounding both these adjusted values was generated as described above with 1000 bootstrap replicates.

### Blood-meal host determination: human vs. animal blood

Mosquito host feeding sources were determined through a qPCR high resolution melt-curve analysis targeting cytochrome B gene fragment, following published protocols.^24^ Briefly, DNA was extracted from 5ul of the above mosquito slurry by combining with 20ul of QuickExtract solution (Lucigen), and incubated for 65°C at 15min with a final inactivation of 98°C for 2 minutes. From this, 2ul of extract was combined with 5ul SsoAdvanced Universal SYBR Green Supermix, 2ul water, and 1ul of 10uM CytB primers, and analyzed with the published amplification/melt conditions in a Mic qPCR Cycler (Bio Molecular Systems, Australia). Bloodmeal discrimination for the purposes of this paper were classified as human if the melting temperature of the amplified product fell between 85.75°C +/- 2°C, and otherwise considered non-human if melt rate (-dF/dT) peaks fell outside this range.

## Results

### Pre-pandemic mosquito background reactivity

Due to the variable nature of mosquito bloodmeal size, a potential range of digestion time per individual mosquito, and high rate of pre-pandemic background reactivity presented in other serological studies,^3,4^ we developed assay cutoffs based on background reactivity from silica gel stored, pre-pandemic, blood-fed mosquito samples (*n=*90). Median Fluorescent Intensity (MFI) values per antigen were non-normally distributed (Shapiro-Wilk normality test, *p-value* < 0.001 for all antigens).

Manufacturer suggested MFI cutoffs for 1:100 dilution of serum (N:450, RBD:250, S1:250, S2:750), were adjusted for this assay using the mean + 5 standard deviations of pre-pandemic mosquitoes to maximize detectability and limit multi-antigen false positivity. These cutoffs (N:116.7, RBD:48.3, S1:78.0, S2:89.4) showed low single antigen positivity (*n*=2, 2.2%) in these mosquitoes during the pre-pandemic period (Figure 2 and Supplemental table 1). Due to this low level of single-antigen positivity and that no pre-pandemic mosquitoes were seropositive based on 2 or more antigens; we chose to classify a mosquito positive only if two or more antigens crossed their specific thresholds.

**Figure 2:**
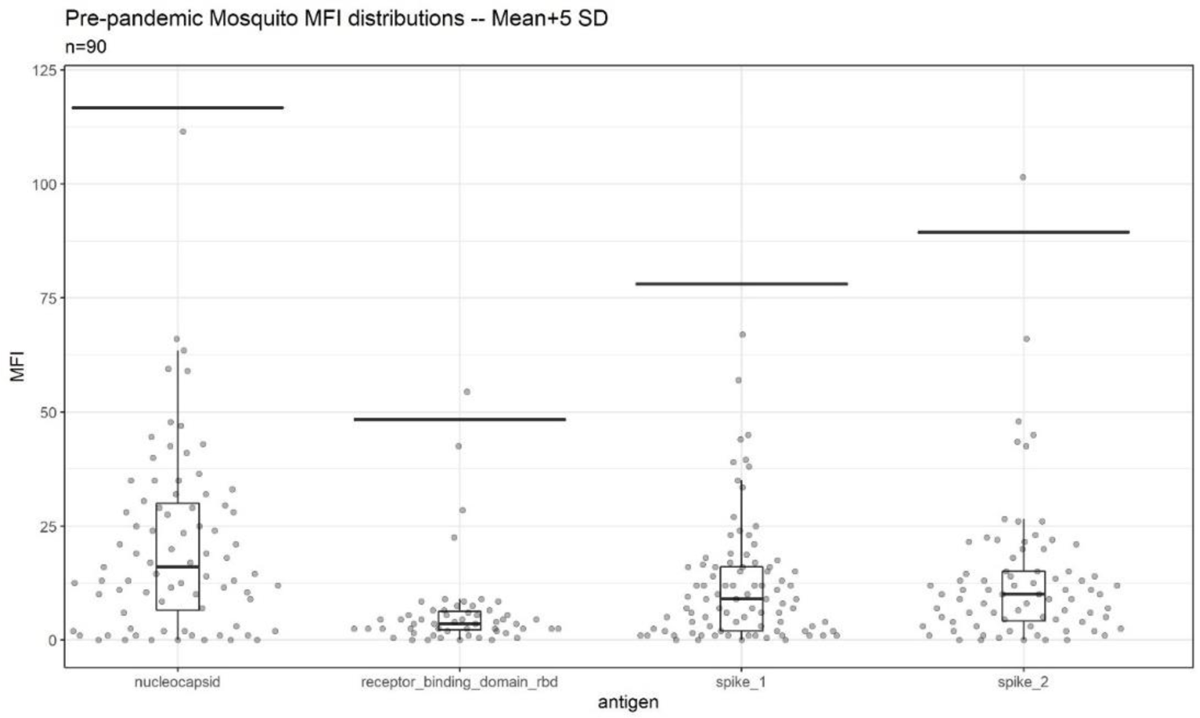
Background reactivity of 90 blood-fed mosquitoes collected via indoor aspiration prior to SARS-CoV-2 emergence and stored desiccated on silica gel until analysis. Per antigen cutoffs are marked via line. The two points above cutoff were single antigen positive mosquitoes

### Durability of antibody-detectability in mosquito midguts

In an initial proof-of-concept time-course experiment, mosquitoes were held for set time points after blood-feeding to assess the detectability of the antibodies in the blood meal during mosquito digestion under normal insectary conditions. Consistent with previous studies on antibodies against other pathogens^13–16, 25, 26^, antibodies against *SARS-CoV-2* remained detectable at least to 10 hours, with a loss of signal seen between that timepoint and 24 hours later (data not shown). Following this pilot, a larger trial was then undertaken with 13 volunteers of unknown SARS-CoV-2 infection history. One volunteer (VAT) had been vaccinated three months prior to the experiment (with an additional suspected infection with SARS-CoV-2 based on high titer against the nucleocapsid antigen, below). VAT had a clear antibody response across all time-points analyzed with 3 or more antigens positive in every mosquito held up to 30 hours post blood-feeding (Supplemental Figure 3). Other volunteers with unknown status were classified based on the positivity rate of mosquitoes in time points 0, 5, and 10 hours, where overall antibody positivity rates were highest (Figure 3). VBT and VDT had all mosquitoes positive with 2 or more antigens through these time points (11/11 and 11/12 mosquitoes, respectively). VFT, VIT, VLT, and VMT each had all but one mosquito positive in these times post-feeding (10/11, 9/10, 4/5, 9/10 total positive mosquitoes, respectively). VGT had two mosquitoes as negative (8/10 positive). These 8 individuals were assumed then to be true positives, with negative mosquitoes being false negatives. On the other hand, volunteers VHT, VCT, VET, and VJT were considered negative due to only 1 or fewer antigen positive mosquitoes at any time point. VKT was also considered negative due to 11/12 mosquitoes having <2 antigens positive but does have one 2-antigen positive mosquito at the 10 hours time-point (Figure 3), indicating this is a false positive (or as an individual with antibody levels very near the limit of detection). Accordingly, misclassification varied between antigens (in time points 0, 5, and 10 hours), being highest in nucleocapsid with 12% false positive and 34% false negative (N=132 mosquitoes), followed by spike1 (2% false positive and 30% false negative, N=132), RBD (0% false positive and 17% false negative, N=132), and spike2 (10% false positive and 6% false negative, N=132). Although the diagnostic values appear to vary among antigens, the small sample size of volunteers available in the present study may have confounded these estimates. Thus, using reactivity against multiple antigens for a diagnosis of a suspected sero-positive is needed to overcome these rates of misclassification based on a single antigen.

**Figure 3:**
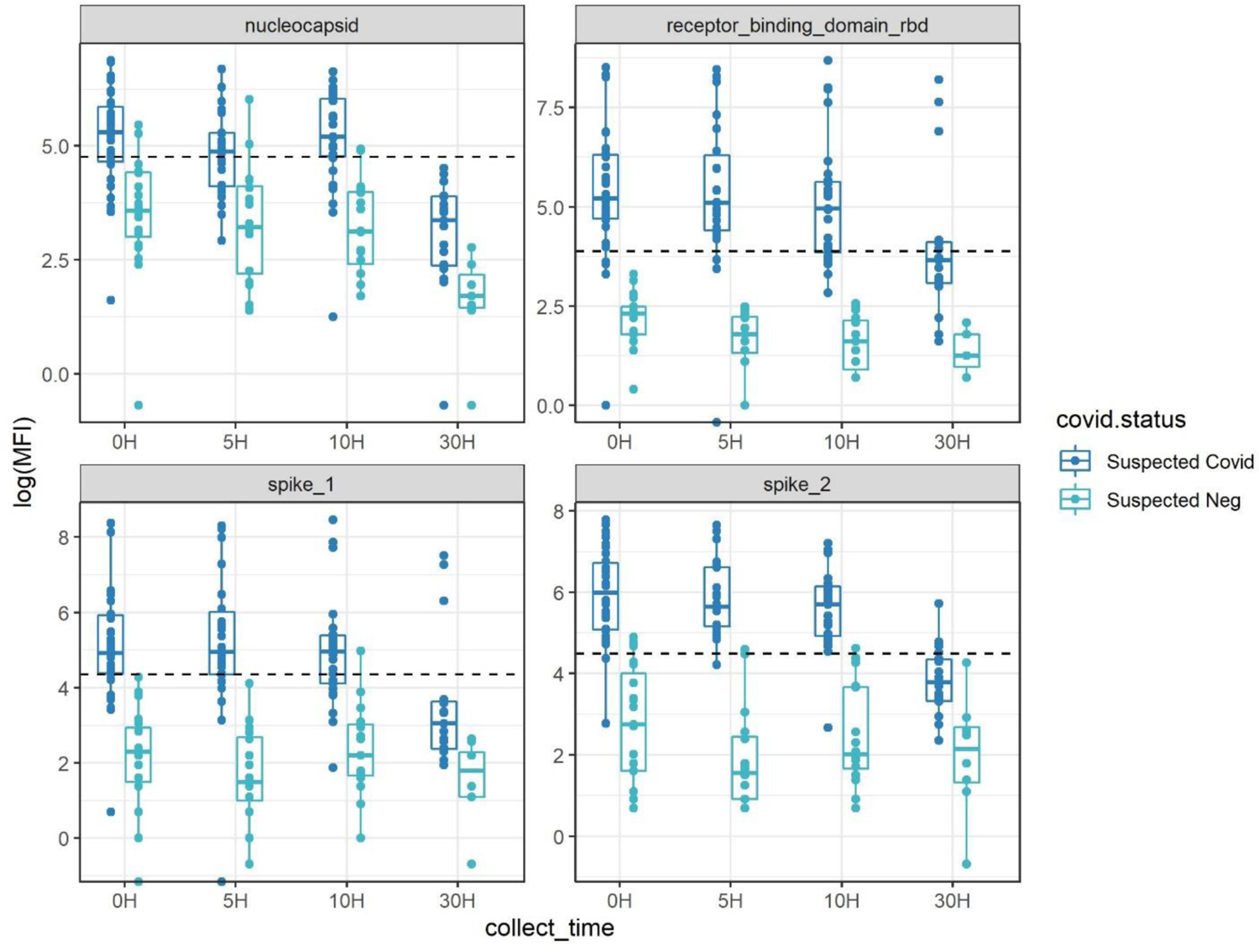
Antibody detectability above antigen-specific cutoffs (dotted line) with the four-antigen multiplex immunoassay after set periods of digestion post-bloodfeeding on human volunteers.

Suspected Covid had 2+ antigens over pre-pandemic cutoffs at multiple time points, suspected negative had a maximum of one antigen positive at any time point. Inconclusive (based on a single mosquito blood meal) had 1 mosquito with 2 antigens positives at various time points (See also Supplemental Figure 1 and Supplemental Figure 2).

Based on these classifications of volunteer infection status and the requirement that reactivity above cutoff must be observed for two or more antigens, we estimated the sensitivity and specificity of our assay throughout these time points using bootstrapping to determine confidence intervals (Table 1).

**Table 1:**
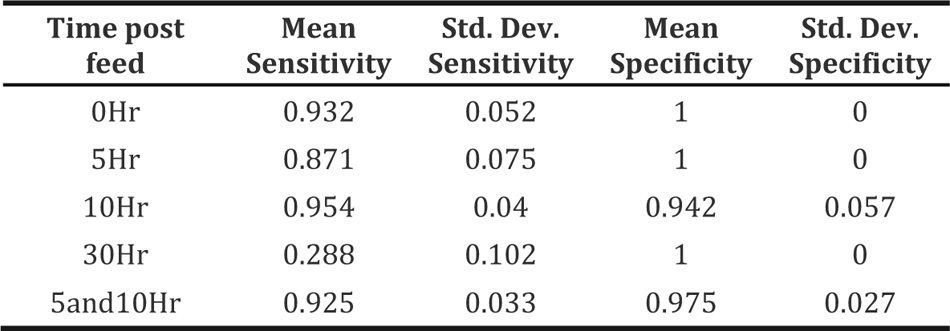
Sensitivity and specificity estimations during each time-point post feeding a total of 159 mosquitoes on 13 volunteers (Figures3 and S1). Standard deviations were calculated using bootstrapping. A 5 and 10 hour merged time-point estimate (5 and 10Hr) was calculated as these are the more likely time-points for collection of blood-fed wild mosquitoes post-feeding than 0 or 30 hours.

| Time post feed | Mean Sensitivity | Std. Dev. Sensitivity | Mean Specificity | Std. Dev. Specificity |
| --- | --- | --- | --- | --- |
| 0Hr | 0.932 | 0.052 | 1 | 0 |
| 5Hr | 0.871 | 0.075 | 1 | 0 |
| 10Hr | 0.954 | 0.04 | 0.942 | 0.057 |
| 30Hr | 0.288 | 0.102 | 1 | 0 |
| 5and10Hr | 0.925 | 0.033 | 0.975 | 0.027 |

We find that while sensitivity is high in 0-10 hours timepoints (0.871-0.954), it drops at 30 hours to 0.288, with only mosquitoes from two of the previously infected volunteers with the highest overall MFI values showing positivity at this time point (Supplemental Figure 2).

### Sero-surveillance of Malian communities

A total of 579 blood-fed mosquitoes (252 were *An. gambiae* s.l. and 327 were *Culex* spp.) collected indoors in five Malian communities by aspiration were subjected to analysis. Numbers of analyzed mosquitoes were randomly subsampled to be largely consistent between sampling periods (284 in October—November 2020, 295 in February 2021), villages (118, 108, 115, 118, 120 mosquitoes analyzed from Bancoumana, Berian, Nionina, Sitokoto, and Sotuba, respectively), and total houses sampled per village (44, 35, 35, 34, 33 in Bancoumana, Berian, Nionina, Sitokoto, and Sotuba, respectively).

An increase in reactivity from the pre-pandemic baseline was apparent across all four SARS-CoV-2 antigens (Figure4). Because we suspected that only a fraction of the population would be seropositive, we used quantile regression to evaluate which quantiles have changed and if the change was consistent across quantiles. An advantage of this approach is that it does not assume cutoffs (Methods), yet it can be used to estimate the quantiles of the population that exhibits crude reactivity changes over the pandemic for each antigen. Considering nucleocapsid and spike-1, an increase over the pre-pandemic was significant at October-November 2020, starting from the 80^th^ and 75^th^ quantiles, respectively (quantile regression, t_df=1_=2.74 and 2.14, P=0.006 and P=0.033, respectively) and increasing in significance at higher quantiles, whereas at Feb-Mar 2021 a significant increase was detected from the 65^th^ and 70^th^ quantiles, respectively (quantile regression, t_df=1_=2.71 and 2.49, P=0.007 and P=0.013, respectively, Figure4b). In RBD, an increase over the pre-pandemic was significant at October-November 2020, starting from the 50^th^ quantile (t_df=1_=3.17 P=0.002) and increasing in significance at higher quantiles, whereas at Feb-Mar 2021 a significant increase was detected from the 30^th^ quantile (t_df=1_=2.89 P=0.035, Figure4b) and increased thereafter. In spike-2, an increase over the pre-pandemic was significant at October-November 2020, starting from the 70^th^ quantile (t_df=1_=2.53 P=0.012) and increasing in significance at higher quantiles, whereas at Feb-Mar 2021 a significant increase was detected from the 50^th^ quantile (t_df=1_=2.43 P=0.016, Figure4b) and increased thereafter. These results, at the single antigen level, exhibited relative change over time, and thus, support a consistent increase from the pre-pandemic baseline. On average across antigens and the five communities, these estimates support 31% crude sero-prevalence in October-November 2020 that increased to 46% in Feb-Mar 2021. Moreover, the increase in the magnitude of the reactivity (Figure4) indicates that a fraction of the population has experienced multiple exposure events resulting in elevated titers among sero-positives (aside from the greater fraction of the population showing an increase over the pre-pandemic levels).

The crude daily rate of infection estimated by the difference in mean prevalence (across antigens in the whole population) between time points divided by the median number of days between samples was 0.13%/d between October-November 2020 and February 2021. Assuming that COVID-19 started spreading in the country one week before the discovery of the first case(s) in Mali (March 25, 2020: http://www.xinhuanet.com/english/2020-03/25/c_138916218.htm), the crude daily infection rate between this and the October sample was 0.15%/d.

Assuming each blood fed mosquito fed on a randomly selected person in the community, crude seroprevalence was estimated based the number of bloodfed mosquitoes with reactivity above cutoff for two or more SARS Cov-2 antigens (above) over the total tested, suggesting a significant increase in four of the five communities between October-November 2020 and February 2021 (Figures5A, S3). Notably, no change or even a trend showing a loss of sero-positivity was recorded for the village Sitokoto, which had very minimal sero-prevalence (1.8%) in October-November 2020 (Figure 5A).

The crude serological data indicates marked heterogeneity over time as well as in space (Table 2). Considering a house “positive” if it had at least one positive mosquito, seroprevalence at the house level was typically, higher than that at the mosquito level, yet the differences over time and across villages were consistent as were the statistically significant differences between periods and across villages (Table 2). Significant differences between villages were detected only at the February 2021 period (Table 2). At the range of seroprevalence measured, there was consistent relationship with house seroprevalence 50% higher than that at the mosquito level (Supplementary Figure 4, Discussion).

**Table 2.**
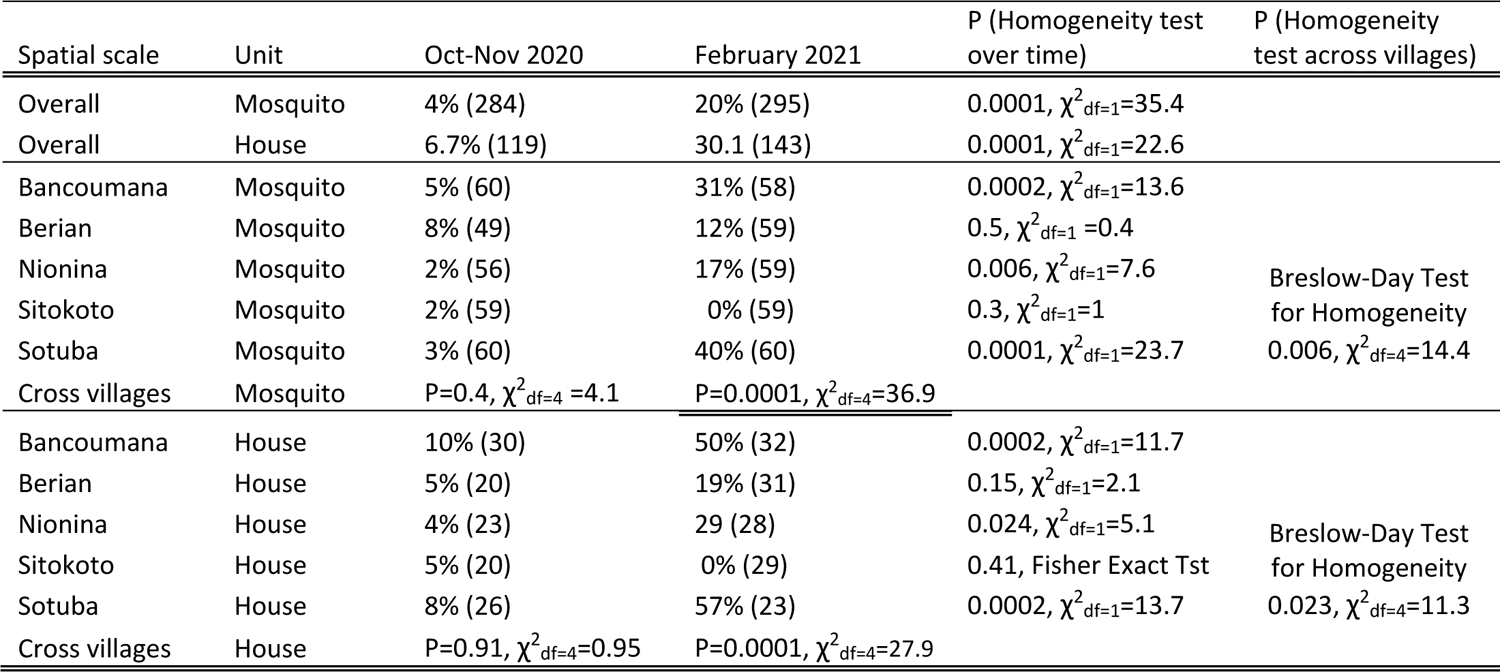
Crude seroprevalence (N) over time across spatial scales and sampling units.

Although these indoor resting mosquitoes are known to feed predominantly on humans ^11, 27–30^ we assessed the variation among villages and time points in this trait, which could confound our results because our secondary antibody was anti-human IgG (Methods). Blood meal analysis to identify human and nonhuman hosts (Methods) was performed on 221 mosquitoes. Overall, 88% fed on human blood including 9% which fed on human and other animal blood (mixed, Supplementary Figure 5). The overall human feeding rate was lower in *Anopheles* (79%, N=109) than in *Culex* (97%, N=112, P=0.001 χ^2^_df=1_ =18.1), was similar between time points (86% in OctNov2020 vs. 91% in February 2021, χ^2^_df=1_ = 1.83, P=0.17) and varied between 66% (Sitokoto) and 97% (Sotuba) among villages (χ^2^_df=4_ = 27.3, P=0.0001, Supplementary Figure 4). With the other villages blood-feeding rate on humans equal or above 91% (χ^2^_df=4_ = 1.6, P=0.6), only the mosquitoes from Sitokoto exhibited an exceptionally low human feeding rate. To accommodate variation in blood feeding rate on our seroprevalence rates, we adjusted the seroprevalence data in each village and time point to the fraction of mosquitoes that fed on humans (Figure5B).

Numbers of mosquitoes collected and analyzed per house across the five villages in each time period varied (Medians=2 and 3, Maximum=11 and 49, respectively, Supplementary Figure 6), and thus we calculated human seroprevalence per village using bootstrap subsampling of one mosquito per house per village per time period (Figure 5C). This led to similar point estimates of prevalence, though with a wider standard deviation.

## Discussion

This is the first study evaluating the use of serological data derived from blood-fed mosquitoes to measure the spread of a non-vector borne disease, namely COVID-19, at a country scale. This approach has a high potential to fill the gap where capacity to effectively sample the target human (host) population directly is low, but where mosquitoes that feed on people are abundant – settings that are common in many developing countries. As this was a proof-of-concept evaluation of this approach, rather than a full-scale investigation (in preparation), we have limited the number of communities, time periods, and samples analyzed. Yet, the results reveal a sharp increase in exposure to SARS-CoV-2 between October 2020 and February 2021, albeit not across all communities. Furthermore, a comparison of key patterns detected here with those established using the classic sero-surveillance study in some of the same Malian communities^8^ suggests high congruency (below). Overall, our results demonstrate that this approach provides valuable insights as to the magnitude of human exposure and its variation over space and time, which can inform epidemiological assessments and decisions.

However, this approach does not convey individual patient exposure status (or seroconversion in repeated sampling) because the individual person the blood came from remains unknown as is the information about their age, gender, etc. Uncertainties regarding the exact volume of the sera a mosquito imbibes, the exact time since blood-feeding, and especially whether a mosquito fed on a human or animal host, and the fraction of mosquitoes that fed on the same people preclude interpreting “mosquito seroprevalence” as identical to the human population’s seroprevalence, without accommodating additional information. Finally, the small volume of blood available in a mosquito (typically 1-5 ul^12^) limits the number of serological assays that can be performed on a single sample. Below, we consider these factors in the analysis and interpretation of the results on the spread of SARS-CoV-2 in Mali and in similar application of this approach in the future for this or other diseases.

Prerequisites for using blood-fed mosquitoes for serological studies include establishing the dynamics of antibody detection over time since blood feeding (using the same preservation method and conditions used in the field) and reactivity cutoffs that are validated using direct feed on local volunteers or blood from seropositive and seronegative individuals from the target populations^6–8^. Early experiments to evaluate the effect of time post feeding on antibody detection revealed that mosquitoes preserved in 80% ethanol indicated rapid reactivity degradation compared with those desiccated on silica gel (not shown). Both laboratory experiments in NIH and field studies in Mali confirmed earlier studies^13, 15, 16, 25, 26^ that antibody detection persisted with minimal degradation until at least 10 hours and degradation was evident at later time points (24-36 hours post feeding, e.g., Figure3). Since most *Anopheles spp.* and *Culex spp.* bite late at night: 22:00 to 04:00^29, 31–35^ and mosquito collection took place from 07:00 to 10:00, most mosquitoes were killed and preserved 3-11 hours post feeding. Moreover, we separated freshly fed mosquitoes which were subjected to serological analysis from later stages of blood digestion including semi-gravid and gravid or non-fed mosquitoes. We established reactivity cutoffs per antigen with wide margin based on the mean and 5 standard deviations, using pre-pandemic, silica gel stored blood-fed mosquito samples from the target population that represent natural background reactivity (Figure2). The low single antigen positivity (2.2%) in the pre-pandemic mosquitoes (Figure2 and Supplemental Table 1) was further minimized by requiring that seropositive mosquitoes exhibit reactivity above cutoffs in two or more antigens. These cutoffs were tested in a trial with 13 volunteers, whose infection history with SARS-CoV-2 was unknown (except for one). Based on highly consistent sero-positivity of the mosquitoes that fed on them (mosquitoes/volunteer >10), the volunteers were readily classified into putative positive and negative states using high consensus among mosquitoes (in time points 0, 5, and 10 hrs Figures3, S1 and S2). Likewise, despite moderate misclassification by single antigens (up to 12% false positive and 34% false negative among *N*=132 mosquitoes in NC)^8, 36^, considering the two-antigen definition at the 5 and 10 hours post feeding time point (above), only 2.5% were false positive mosquitoes and only 7.5% were false negative mosquitoes, assuming the classification of individuals was correct (Table 1).

Overall, results based on 669 blood-fed mosquitoes collected indoors across five Malian communities (Bancoumana, Berian, Nionina, Sitokoto, and Sotuba following collection in two pre-pandemic villages) revealed an increase in reactivity from the pre-pandemic baseline across all four SARS-CoV-2 antigens (Figure 4). This increase was significant between the pre-pandemic and the early (October-November 2020) and late (February 2021) pandemic time periods (Figures 4 and 5), but also between the early and late pandemic time periods (quantile regressions, P<0.01 and Table 2). Assuming minimal change in confounders such as human feeding rate, this trend presents a compelling proof for the utility of mosquito-based analysis of disease spread especially because it does not depend on cutoff values. This analysis indicated a steady increase of the fraction of the population exhibiting elevated reactivity over the pre-pandemic level as well as elevated intensity of the reactivity across the higher quantiles (Figure4), suggesting higher titers among putative positives, as expected if people are repeatedly infected when more individuals carry the virus. Assuming these five, mostly rural communities represent the whole of Mali, the crude daily rate of infection (estimated by the difference in mean prevalence across antigens between time points divided by the median number of days between samples as explained above) was 0.13%/d between October-November 2020 and February 2021. Assuming that COVID-19 started spreading in the country one week before the discovery of the first case(s) in Mali (above), the crude daily infection rate between this and the October sample was 0.15%/d. Albeit lower than those reported from Mali^8^, the difference may reflect the more remote and rural settings of the communities sampled here. Indeed, at Doneguebougou, the most rural community sampled by Sagara et al. (2021^10^, which is located ∼15 km from Bamako, their estimate for the same time period was similar (0.19%/d), and unlike our estimate, their rate included persons who were positive at the first time point and negative in the second time point.

**Figure 4:**
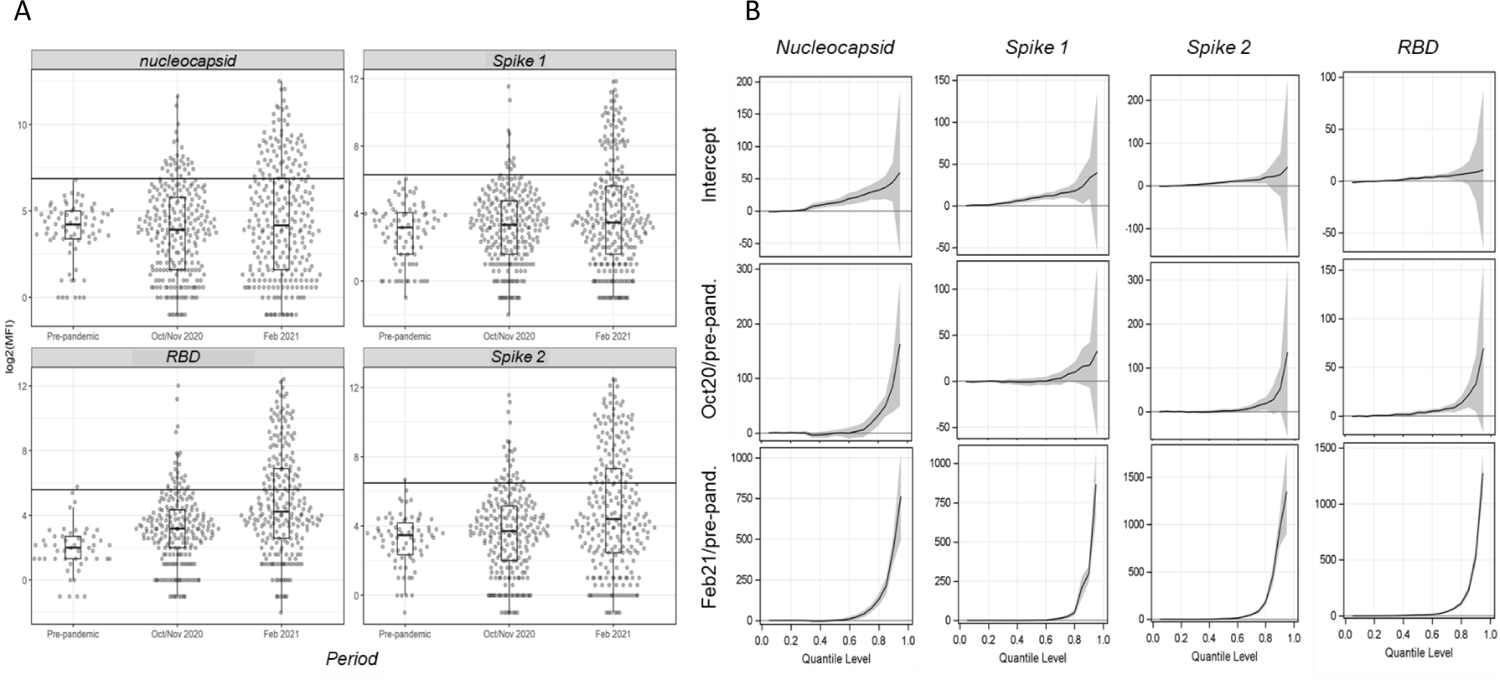
Distributions of the median fluorescence intensity (MFI) per antigen comparing pre-pandemic (N=90) and pandemic mosquitoes (October-November 2020, N=284; and February 2021, N=295) and quantile regression results showing quantile specific changes in reactivity over time for each antigen. (a) Reactivity distribution of each antigen and time period overlaid with box-whisker plots. The cutoffs are shown by the horizonal lines. (b) Results of quantile regression models fitted to each antigen with period as the independent variable, showing the intercept and the effect of each pandemic time period relative to the pre-pandemic baseline with 95% confident interval (blue band). Line segments above zero indicate quantiles in which the effect is positive and statistical significance is indicated if the CI range does not overlap with the zero baseline.

Following the definition of sero-positive mosquito’s bloodmeal (reactivity > cutoff in two or more SARS Cov-2 antigens), we estimated the crude population seroprevalence of each community and time point, assuming each mosquito fed on a randomly selected resident (Figures 5 and S3). That the seroprevalence at the house level was 50% higher than that at the mosquito level (Table 2, Supplementary Figure 4) reflecting the combined effects of the clustering of seropositive persons between houses in a village, the number of mosquitoes analyzed per house, and the fraction mosquitoes that blood fed in one house overnight and moved into another by morning^37, 38^. Because of this and the quicker saturation of the house seroprevalence (defined as having at least one seropositive mosquito in a house at a given time period), we suggest that the crude seropositivity at the mosquito level provides more accurate estimate of the community true seroprevalence. Additionally, blood-fed mosquitoes should be sampled from at least 25 houses in the community, and possibly from a larger number based on its total size, spatial organization, and heterogeneity with respect to relevant factors, e.g., proximity to school, market, etc. Overall, the crude seroprevalence rate at October-November 2020 was 6.5% (Sitokoto: 1.8%—Berian: 12.2%, Figure5A, Table 2), representing seven months after the discovery of the first case of COVID-19 in Mali. However, three and a half months later (February 2021), the overall crude seroprevalence was dramatically higher: 25.0% (Sitokoto: 0%—Sotuba: 46.5%, Figure5A, Table 2). This rise corresponds to the first peak of elevated transmission in Mali (November 2020—January 2021, Figure1).

**Figure 5:**
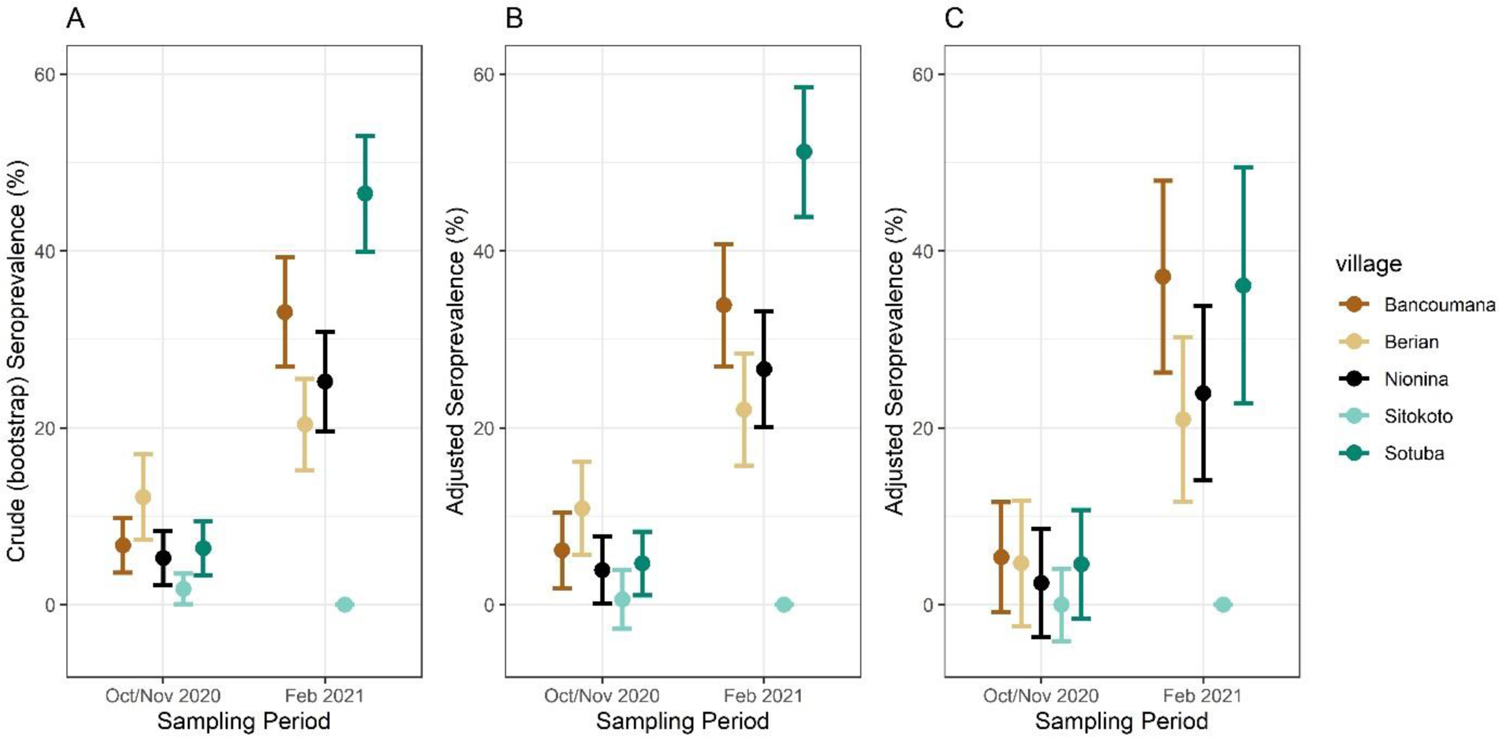
Crude (A) and adjusted seroprevalence per sampling village. Adjusted seroprevalence for test sensitivity and bloodmeal composition (B), and test sensitivity with one mosquito per sampling period per household (C).

Our crude seroprevalence may underestimate actual human population seroprevalence because the assay’s sensitivity was lower than its specificity (Table 1) while the majority of the population would be still sero-negative and because some of the mosquitoes had taken their blood meal on non-human hosts (which our ELISA cannot detect even if that host had antibodies against SARS-Cov-2). The adjusted seroprevalence values were typically 2% higher than the crude seroprevalence across communities and in each one, except in Sotuba during February 2021, where the adjusted seroprevalence was 4.5% higher than the crude value (Figure 5). Overall, 12% mosquitoes fed on non-human blood (N=221), aside from 9% that fed on human and other animal blood, proportions that are consistent with previous studies^11, 27, 29, 32, 33^. None of the mosquitoes that fed on animal blood were seropositive (N=26; 1 mosquito was reactive to a single antigen). Minor difference was detected between October-November 2020 (86%) and February 2021 (91%, above) and feeding on human blood was above 91% in all villages except in Sitokoto (66%), which also had the lowest crude seroprevalence. Incorporating a bead that indicates human IgG or other human-specific antigen into a single ELISA would be helpful in future studies, especially in areas where feeding on non-human hosts is more common. Finally, to consider the possibility that mosquitoes collected in the same house fed on the same person, we also estimated the human seroprevalence by resampling one mosquito from each household (Figure 5C). Because most houses had >3 occupants, and the number of mosquitoes analyzed from the same house at each time point was small (median=2, Supplementary Figure 4), the expected effect of this adjustment was small. A large-scale analysis of sampling of mosquitoes across ∼20 communities in Mali is currently underway, with investigation of ELISA-based techniques better suited to lower resource laboratories to further elucidate the temporal and spatial spread of the virus across the country using this approach.

## Conclusions

The congruence of our results based on serological analysis of blood-fed mosquito with conventional serological studies^8^ and with active infection records based on PCR carried out in Bamako (Fig 1) lend strong support for the utility of this approach. Akin to wastewater-based epidemiology^39^ this non-invasive blood sampling is a promising tool to monitor populations in areas where robust serological data from human subjects is unlikely to be available and where human biting mosquitoes are common, as is the case in many tropical remote communities. While these population-targeted techniques should be thought of as complementary to and distinct from direct serological studies on human populations, they have been proven to be relevant and useful for public health (community-wide) decision making^39, 40^. Understanding exposure rates to pathogens in remote communities as well as changes in reactivity over time are important components of an early warning system targeting remote tropical communities, especially for rare and emerging conditions where conventional surveillance may be considered too costly. Combined with the identification of the blood source, blood-fed mosquito analysis may also be useful to monitor pathogen exposure rates in both human and animal hosts, even if these hosts are poorly characterized (i.e. spillover into an intermediate unknown hosts). Thus, this technique and other future interrogations of the mosquito blood meal could fit well in a one-health paradigm surrounding disease transmission throughout the home or screening across a diverse set of potential reservoirs.

## Data Availability

All data produced in the present study are available upon reasonable request to the authors

## Acknowledgements

We are grateful to the residents of the Malian villages for their permission to work at and near their homes and for their wonderful assistance and hospitality. We thank Drs. Thomas Wellems and Carolina Barillas-Murry, Ms. Wendy Hamm, Ms. Fatoumata Bathily, and Mr. Sam Moretz for the support of our program at NIH and the ICER Mali. This study was supported by the Division of Intramural Research, National Institute of Allergy and Infectious Diseases, National Institutes of Health, Bethesda MD, USA (Grant ID: AI001328)

## Supplementary Materials

**Supplemental Figure 1:**
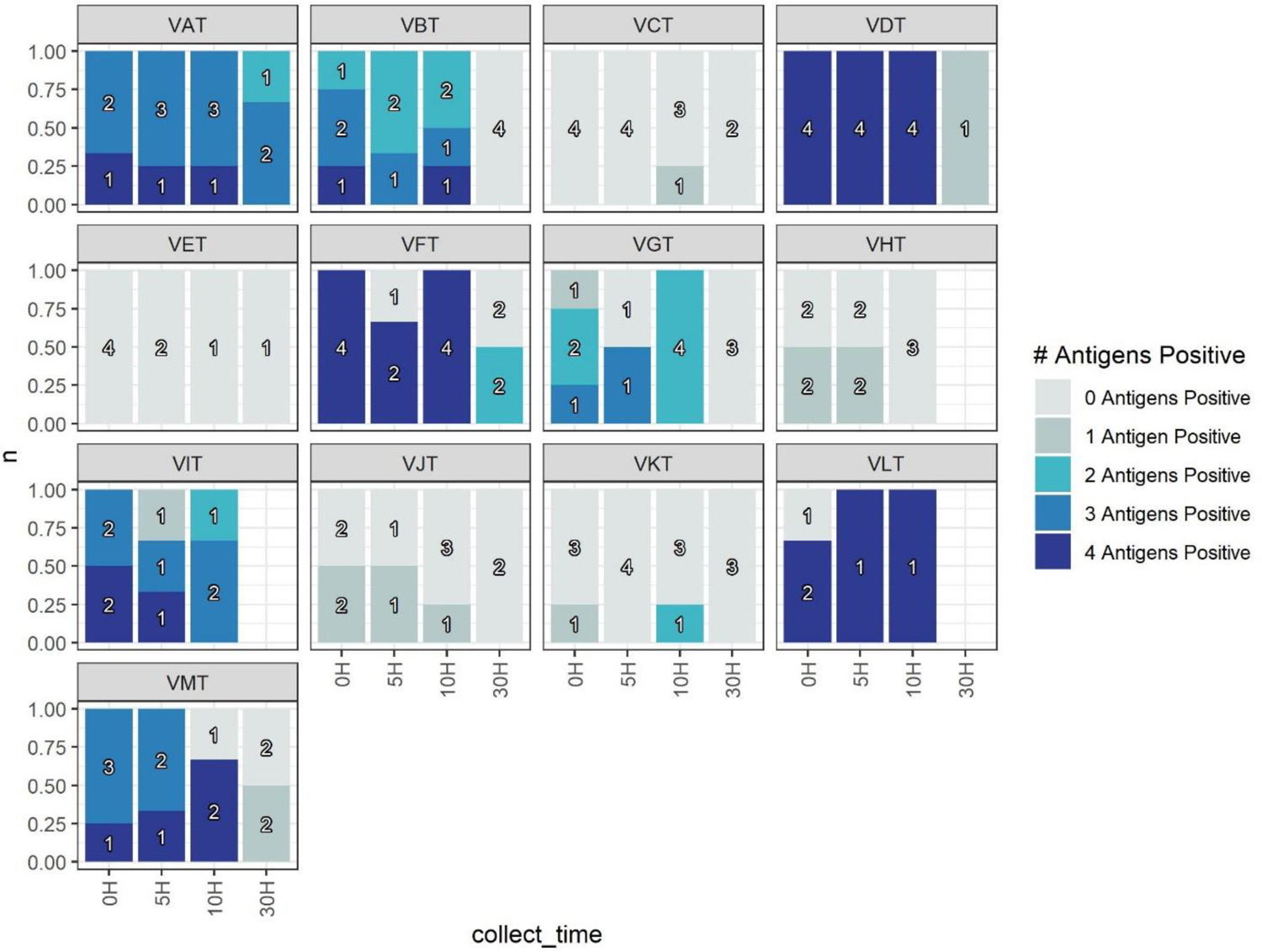
Numbers of mosquitoes positive for 1-4 anti-SARS-CoV-2 antigens per volunteer at 0, 5, 10, and 30 hours post-feed. Only VAT had known covid status with a SARS-CoV-2 vaccination. All other volunteers had unknown SARS-CoV-2 infection history. Per antigen positivity is defined by pre-pandemic collected wild mosquitoes that fed naturally on Malian individuals. Individuals VAT, VBT, VDT, VFT, VGT, VIT, VLT, and VMT are considered positive by having multiple 2+ antigen positive mosquitoes across multiple time points. VHT, VCT, VET, and VJT are considered negative due to only 1 or fewer antigen positive mosquitoes at any time point. VKT is considered negative due to 11/12 mosquitoes having <2 antigens positive, but does have one 2-antigen-positive mosquito at the 10 hours time-point.

**Supplemental Table 1:** Number mosquitoes with the listed number of SARS-CoV-2 antigens above <u>cutoffs per time period.</u>

| Time Period | # Antigens Positive | Total tested |
| --- | --- | --- |
| Pre-pandemic | 0 | 88 |
| Pre-pandemic | 1 | 2 |

**Supplemental figure 2:**
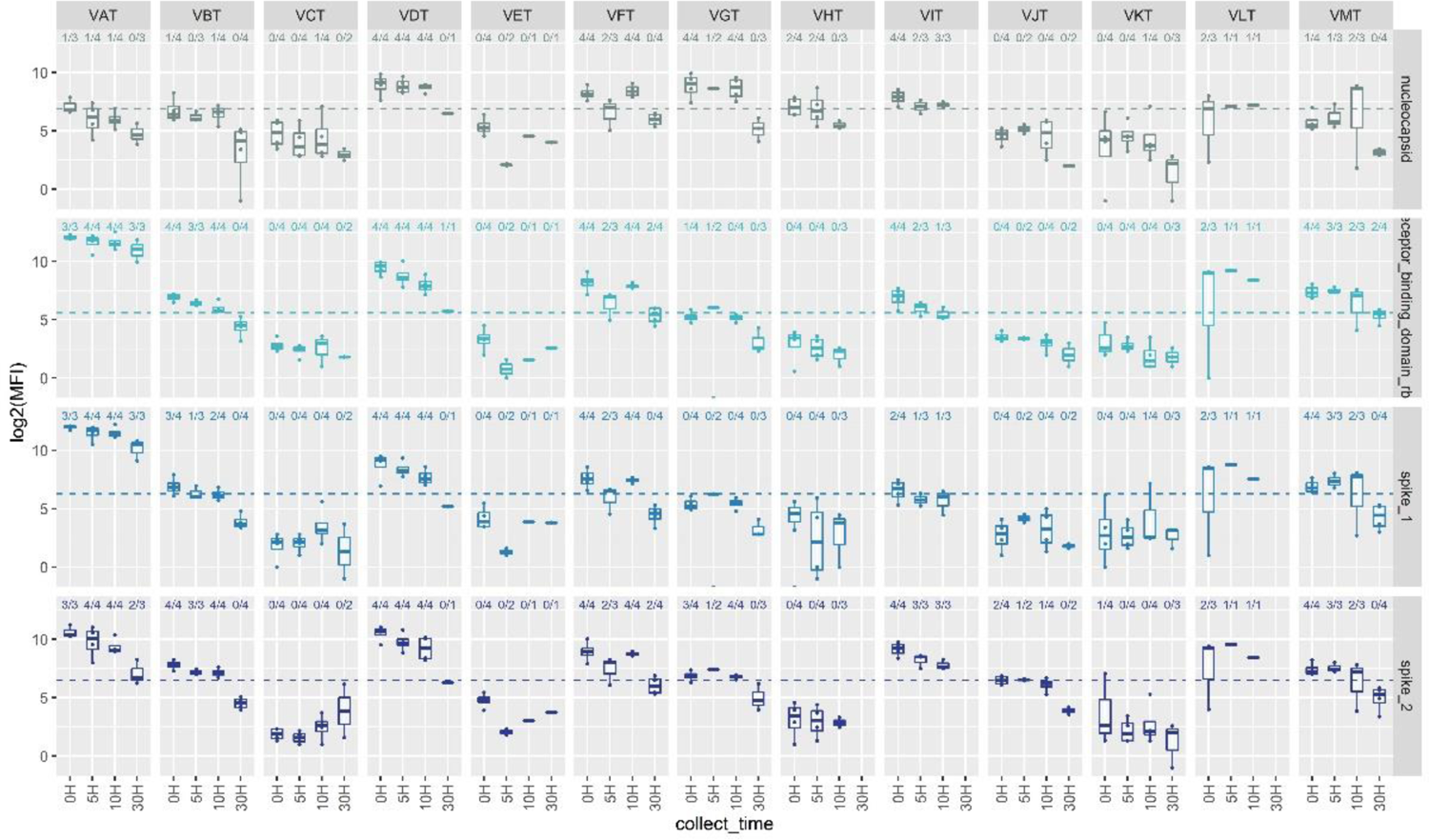
Per antigen log2(MFI) values for each volunteer with pre-pandemic cutoffs shown as dashed line per antigen. Number of mosquitoes per positive per antigen/volunteer per time point labeled above boxplots.

**Supplemental figure 3:**
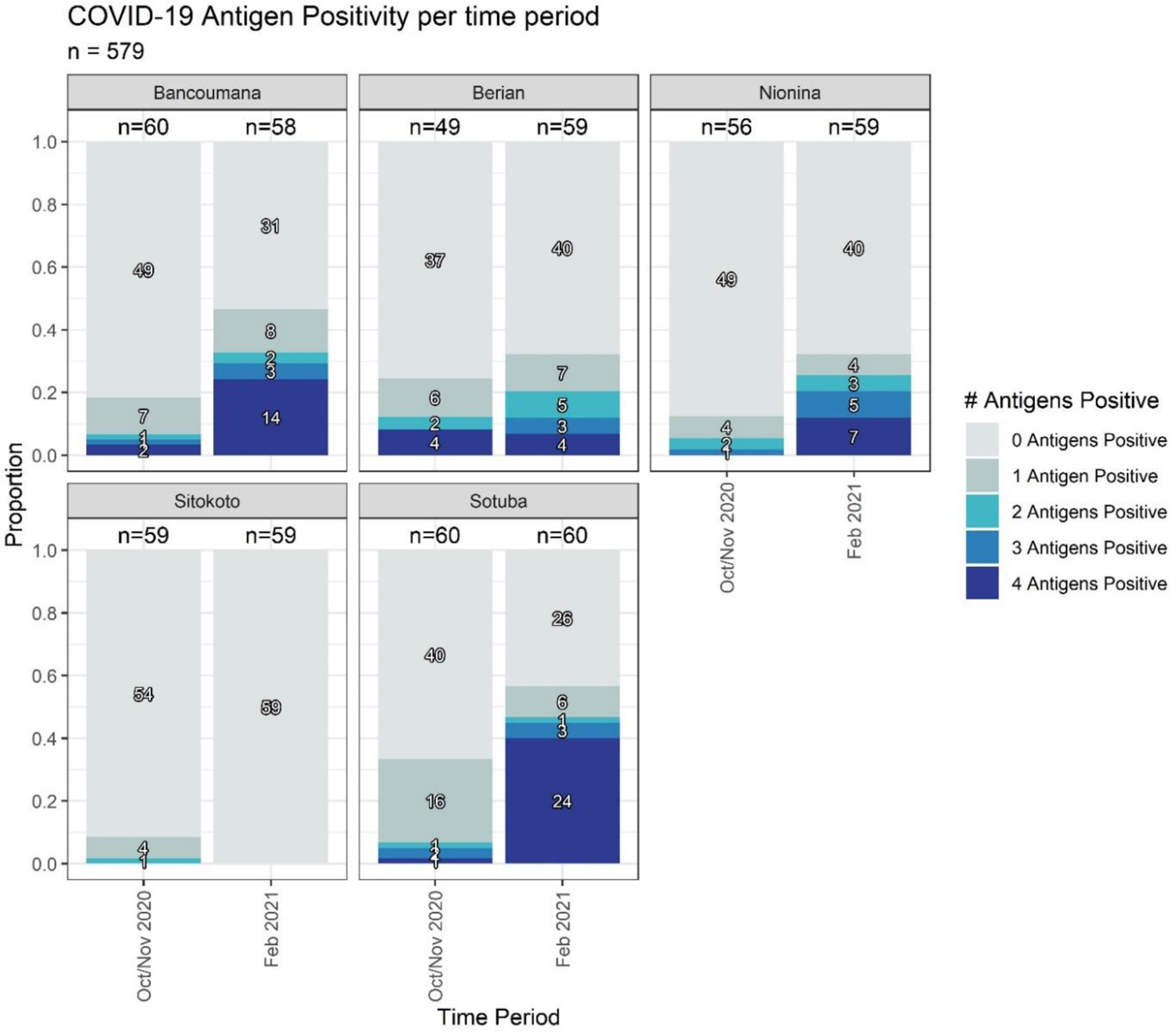
Changes in seropositivity over time measured by the number of antigens to which reactivity exceeded cutoff by village

**Supplemental Figure 4:**
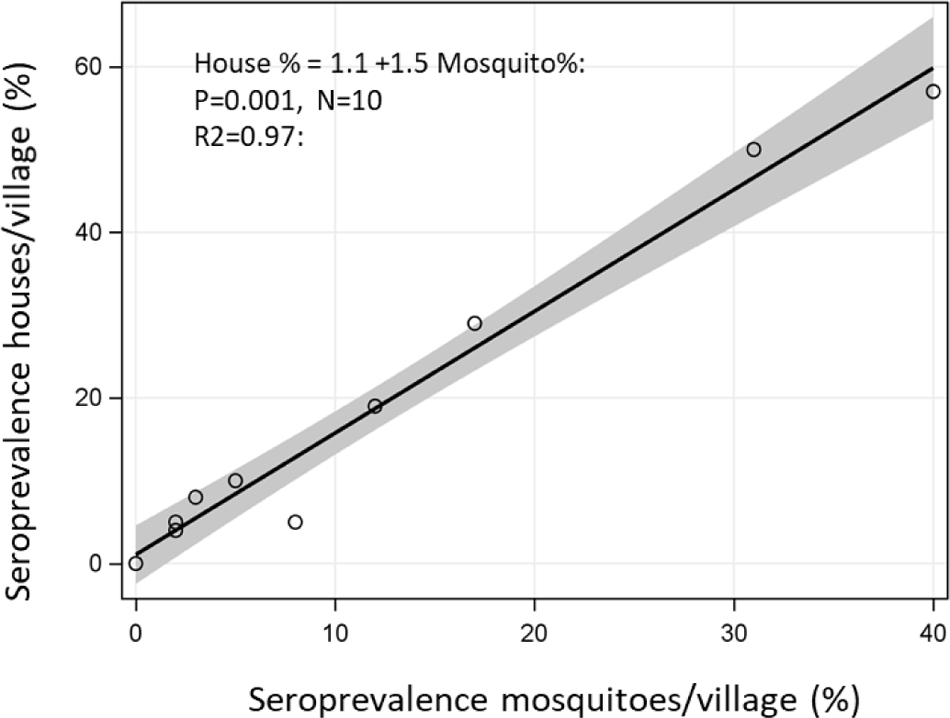
Relationship between seroprevalence at the mosquito and the house levels (across villages and time points).

**Supplemental Figure 5:**
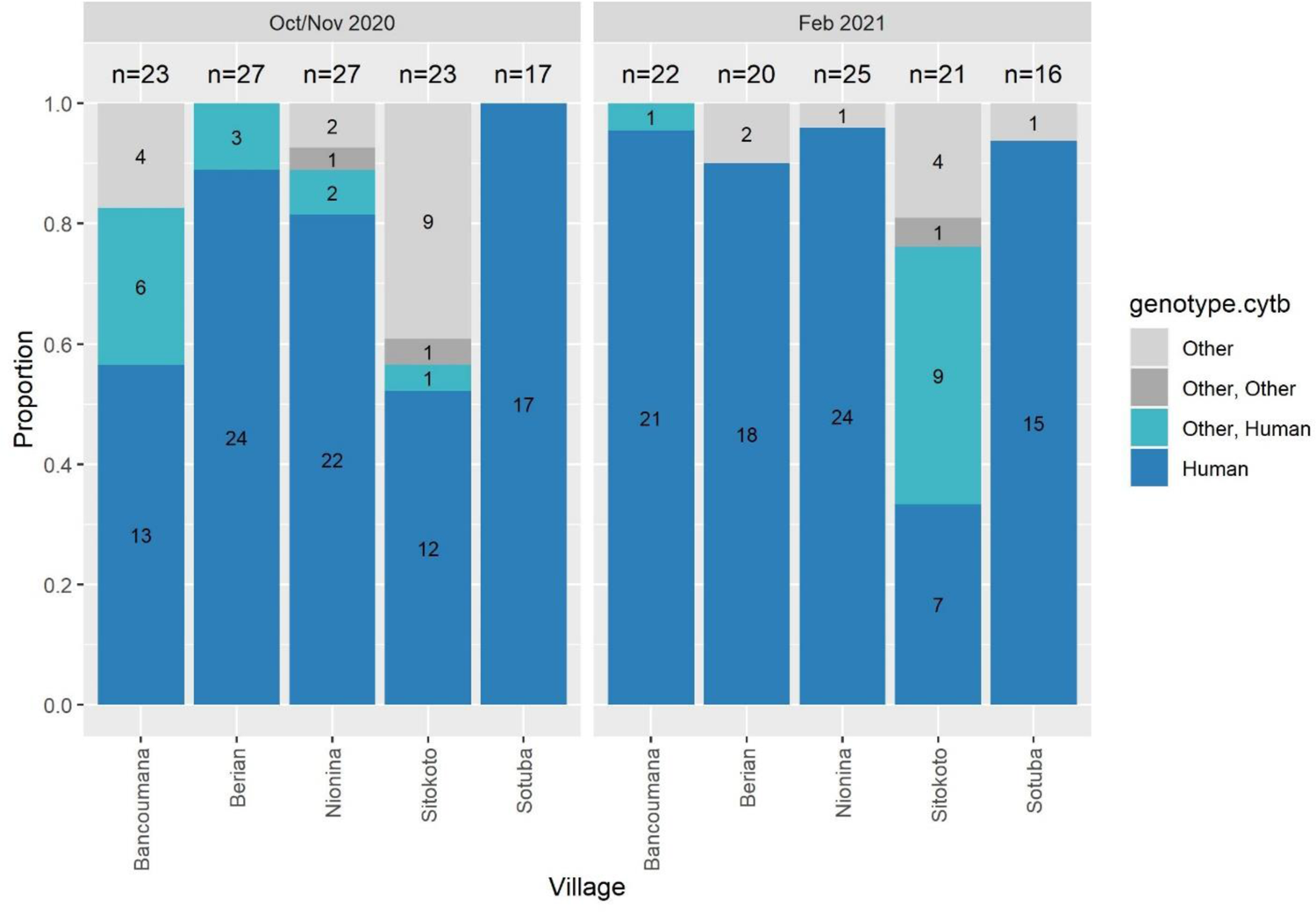
Blood meal composition across villages and time periods (N=221).

**Supplemental figure 6:**
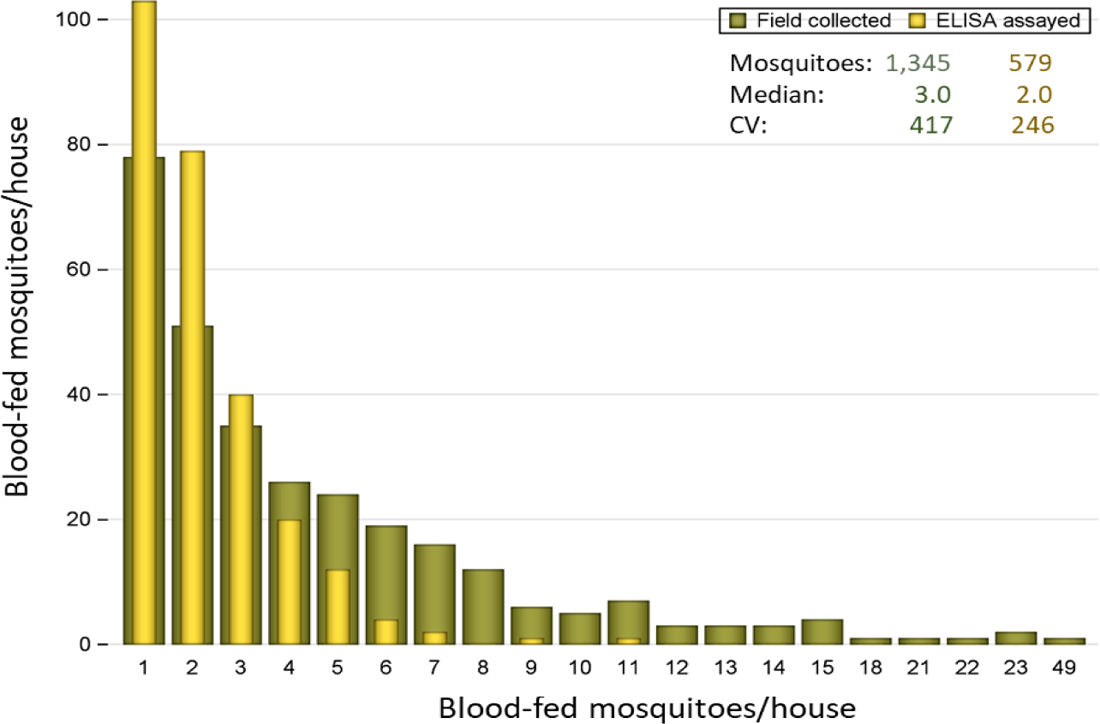
Distribution of the number of mosquitoes collected (green) and analyzed (gold) per house across the five villages by time period (occasionally multiple collection days per time period). Note: the X-axis is not continuous.

